# The Evolutionary Dynamics and Regional Spread of Mpox in Africa: Insights from Multi-country Genomic Surveillance

**DOI:** 10.64898/2026.04.07.26347884

**Authors:** Collins Kipngetich Tanui, Eddy Kinganda Lusamaki, Áine O’Toole, Moses Chitenje, Allan K.O. Campbell, Moussa Moïse Diagne, Stephen Kanyerezi, Martin Faye, Samuel Oluwafunmbi Ifabumuyi, Néhémie Nzoyikorera, Héritier Obed Lango, Félix Koukouikila-Koussounda, Syndou Meite, Elvis Sikazwe, Delia Doreen Djuicy, Bright Adu, Issaka Maman, Lawrence Amon Mapunda, Dougbeh Chris Nyan, Sohou Stephane, Stephen Akuma Aricha, Thibaut Armel Chérif Gnimadi, James Ayei Maror, Amilton Miguel Pereira, Yao Selom Atrah, Olusola Anuoluwapo Akanbi, Emmanuel Lofiko Lokilo, Princesse Tshambu Paku, Jean-Claude Makangara-Cigolo, Gradi Ndelemo Luakanda, Adrienne Amuri-Aziza, Tony Wawina-Bokalanga, Ibrahim Mugerwa, Andrew Nsawotebba, Alisen Ayitewala, Anu Jegede Williams, Vidalyn Folorunso, Sia Mani, Doris Harding, James Christopher Avong, Chimaobi Chukwu, Adama Ahmad, Marie Noelle Uwineza, Dionis Nizigiyimana, Cassien Nduwimana, Theogène Ihorimbere, Boniface Koffi, Ornella Anne Sibiro, Christian Noël Malaka, Pembe Issamou Mayengue, Reiche Golmard Elenga, Dachel Aymard Eyenet Boussam, Vincent J. Munster, Claude Kwe Yinda, Albert Konan Yavo, Edgard Valery Adjogoua, Yakoura Karidja Ouattara, Aboubacar Sylla, Joseph Mutale, Cherry Kasongo, René Ghislain Essomba, Linda Esso, Nadine Lamare Boutgam, Célestin Godwe, Ginette Edoul Mbesse, Salum Nyanga, Jackson Peter Claver, Melak Getu, Bode Ireti Shobayo, Kalilu Samuka Donzo, Sengbe Opati Nyilah, Goundote Aimé, Abdi Roba, Shaline Akasa, Samson Ireri, Millicent Nyakio Ndia, Abdoul Karim Soumah, Haby Diallo, Alpha Kabiné Keita, Abdoulaye Toure, Alex Mwanyongo, Christopher Misomali, Kagoli Matthew, Joseph Daniel Wani Lako, Gregory Wani Dumo, Lul Lojok Deng, Joana Filipa Machado de Morais Afonso, Neto De Vasconcelos, Nuno Rodrigues Faria, Illich Manfred Mombo, Gaël Darren Maganga, Nadine N’dilimabaka, Antoinette Grobbelaar, Jonathan Featherston, Abebaw Kebede, Luzia Lorse Mendes Inglês Samuel, Andrew Ajang Othow, Joseph Bitilinyu Bangoh, Mirriam Nyenje, Alpha Kabinet Keita, Esther Mwau Sigilai, Yadouleton Anges, Gemechu Tadesse Leta, Getachew Tollera, Ambele Mwafulango, Joseph Humphrey Kofi Bonney, Charles Kouanfack, Ahidjo Ayouba, Jacqueline Weyer, Hadja Hamsatou, Roma Chilengi, Herve Alberic Kadjo, Emmanuel Nakoune, Abdourahmane Sow, Fabien Roch Niama, Ernest Lango-yaya, Joseph Nyandwi, Jide Idris, Olajumoke Babatunde, Foday Sahr, Richard Njouom, Susan Nabadda, Andrew Rambaut, Isaac Ssewanyana, Placide Mbala Kingebeni, Sofonias Kifle Tessema, Yenew Kebede Tebeje

## Abstract

The recent MPXV epidemic across Africa revealed extensive viral diversity and complex transmission dynamics, prompting a continent-wide genomic investigation. We analysed 3,450 high-quality MPXV virus whole genomes from 24 African Union Member States, revealing the complex and concurrent circulation of Sub-clades Ia, Ib, IIa, and IIb. Subclade Ia showed high levels of virus diversity in reservoir hosts in Central Africa, detected through zoonotic transmission and some sustained human outbreak lastly detected. In contrast, Clade Ib exhibited signatures of sustained human-to-human transmission across Eastern and Southern Africa. Clade IIa remains largely zoonotic in West Africa. Like Ia, IIb shows continued zoonotic transmission, and sustained human outbreak linked to lineage G1 and G2 circulation. Phylogeographic analyses revealed frequent cross-border transmission and interconnectedness, which was aligned with both human mobility corridors and international boundaries. For instance, the Democratic Republic of the Congo or Sierra Leone seems to emerge as a source of regional exportation, while the Cameroon–Nigeria, CAR-Cameroon or CAR-DRC interfaces reflected ongoing cross-border zoonotic spillovers. These findings underscore the need for harmonised genomic surveillance, APOBEC3-aware triage, and integrated One Health strategies to prevent local outbreaks from escalating into regional epidemics and to inform vaccine deployment and public health preparedness.

## Background

Mpox, formerly known as monkeypox, is a historically zoonotic viral disease caused by the mpox virus (MPXV) in the *Orthopoxvirus* genus. It was first identified in humans in 1970 in the Democratic Republic of Congo (DRC). For decades, it remained largely confined to the rural areas of Central and West Africa, with sporadic outbreaks and limited regional and global attention. Between 2022 and 2025, over 148,000 cases and nearly 1800 deaths were reported across 23 African countries ^1^. The detection of new variants, such as clade Ib in eastern DRC, and the spread to previously unaffected countries like Burundi, Rwanda, Uganda, Kenya and others, underscores the evolving threat. In response, Africa CDC declared a Public Health Emergency of Continental Security (PHECS) in August 2024, followed by the WHO’s declaration of a Public Health Emergency of International Concern (PHEIC) - both signalling the urgent need for strengthened surveillance, diagnostics, and coordinated public health action ^2,3^.

The MPXV epidemic has highlighted a complex and evolving viral landscape across the African continent, shaped by ecological diversity and human mobility, of which there is novel insight given the increasing sophistication of genomic surveillance ^4–6^. Sampling worldwide and across the African continent has described four major clades of MPXV -Ia, Ib, IIa, and IIb ^5^. The number of documented zoonotic MPXV cases has risen sharply in recent years, likely reflecting either a true increase in spillover events or improved detection through enhanced surveillance; nonetheless, MPXV is now recognised as a re-emerging pathogen with epidemic potential ^7^.

While each clade has been linked to sporadic zoonotic infections at different levels, three sustained human-to-human (H2H) outbreaks have been documented and have been named sh2017 (IIb), sh2023 (Ib), and sh2024 (Ia), respectively ^8^. The IIb-associated global outbreak of 2022–2023 (sh2017) spread across continents and exhibited enrichment of APOBEC3-like mutations accumulating in the virus population as it transmitted in humans. In Africa, lineage A within clade IIb exhibited high proportions of APOBEC3-type mutations compared to earlier outbreaks, indicating sustained human-to-human transmission ^9^. Notably, the G1 lineage deriving from A.2.2 was traced from Sierra Leone to Germany and the USA, confirming travel-associated exportation ^10^. The Ib-associated Eastern DRC outbreak in 2023 (sh2023) showed localised but persistent transmission in South-Kivu ^6,7,11^ before spreading to neighbouring countries, including Kenya, Rwanda and Burundi ^7^. Finally, Clade Ia, previously unrecognised diversity in the DRC, suggests an under sampled reservoir population ^12^. Added to the cross-border spread between CAR and DRC, the Clade Ia-associated outbreak in 2024 (sh2024) emerged after repeated introductions in urban settings such as Kinshasa, DRC ^13^.

Broader drivers—urbanisation, cessation of smallpox vaccination, intensified human–wildlife contact, and high population mobility—likely modulate both spillover risk and onward transmission (Wang et al., 2025). The emergence of multiple clades capable of sustained transmission, coupled with evidence of sexual transmission networks, particularly among MSM populations, signals a need for targeted public health interventions ^14,15^.

Several studies have explored the molecular epidemiology of MPXV clade Ia, Ib, IIa, and IIb separately in the endemic and affected countries ^7,13,16–21^. A few of them have provided in one report a detailed, large-scale genomic analysis, transmission dynamics, and over an extended period, specifically focused on most of the countries in Africa. In a recent paper, global genomic surveillance of MPXV (Otieno et al., 2025), about 10,670 MPXV complete sequences from 65 countries collected between 1958 and 2024 have been analyzed. The results reveal high mobility of clade I viruses within Central Africa, sustained H2H transmission of clade IIb lineage A viruses within the Eastern Mediterranean region, and distinct mutational signatures that can distinguish sustained human-to-human from animal-to-animal transmission. The study highlights the importance of genomic surveillance in tracking the spatiotemporal dynamics of MXPV clades. It emphasises the need to enhance such surveillance, particularly in certain parts of Africa.

Here, we present a continent-wide genomic and epidemiological overview of MPXV in Africa, integrating available and high-quality genome sequences from 23 African Union Member States, including data up until 2025-09-01. This dataset not only refines our understanding of clade structure and diversity but also provides critical insights into the dynamics of cross-border transmission. This study contributes not only a pan-continental genomic atlas of MPXV but also a framework for understanding its evolutionary and transmission dynamics. By combining genomics with surveillance data, we not only map the current situation across the continent but also introduce substantial new genomic evidence to refine our understanding and to inform outbreak prediction, diagnostic and vaccine development, and tailored control strategies.

## Results

In an unprecedented continental effort, we gathered 3,450 complete or near-complete MPXV virus (MPXV) genomes from clinical samples, including 1,630 Clade Ia genomes, 1,247 Clade Ib genomes, 34 Clade IIa genomes, and 539 Clade IIb (Figure 1a), which were analyzed with a set a contextual genome previously made available in public repositories. These genomes were collected across 24 African Union (AU) Member States. The AU Member States included Angola, Benin, Burundi, Cameroon, Central African Republic (CAR), Côte d’Ivoire, Democratic Republic of the Congo (DRC), Egypt, Ethiopia, Gabon, Ghana, Guinea, Kenya, Liberia, Malawi, Nigeria, Republic of the Congo (RoC), Sierra Leone, South-Africa, South Sudan, Togo, Uganda, United Republic of Tanzania and Zambia (Figure 1b). This broad geographic representation enabled a nuanced understanding of regional transmission dynamics and evolutionary trajectories. This expansive dataset spanning West, Central, East, Northern, and Southern Africa marks a transformative moment in pathogen genomics, offering the most comprehensive view to date of MPXV Clade diversity and evolution in Africa (Figure 1).

**Figure 1.**
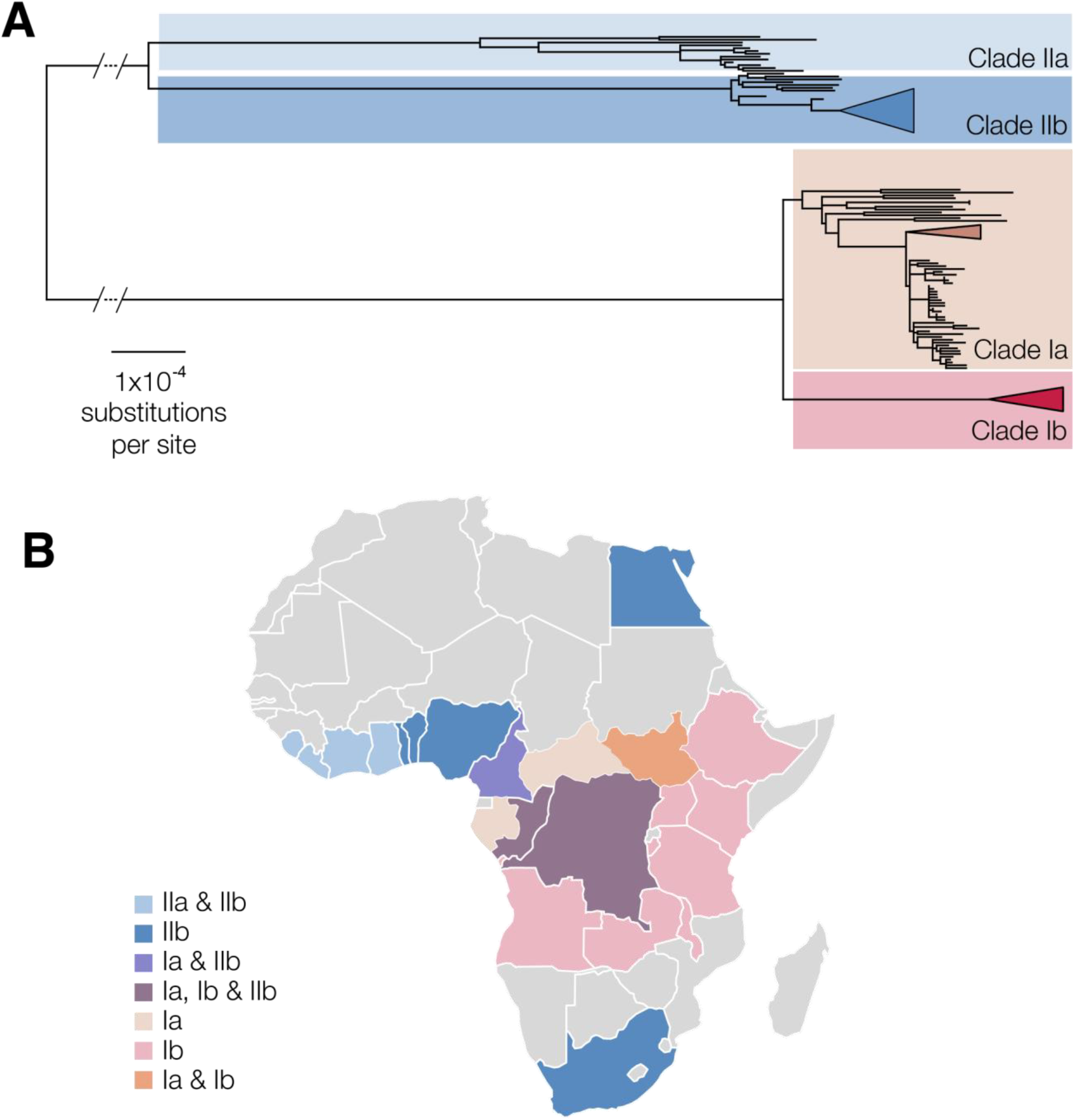
**A)**. The genetic diversity of MPXV is separated into two major clades, Clade I (broadly, Central African) and Clade II (West African). To date, three sustained human outbreaks have been characterised, represented as triangles in the phylogeny and corresponding to sustained human outbreaks (sh)-2017 (clade IIb), sh2023 (clade Ib) and sh2024 (clade Ia). **B). Overall Distribution of MPXV Virus Clades in Africa**. The distribution of MPXV virus clades, including Clade Ia, Ib, IIa and IIb, and their relationship to recent outbreak lineages.

### Genomic diversity of the MPXV Clade Ia

A total of 1,630 MPXV Clade Ia genomes from four Central African countries were analyzed. The dataset was predominantly composed of genomes from the Democratic Republic of Congo (DRC, n = 1,514 [92,88%]), alongside contributions from the Central African Republic (CAR, n = 92[5.64%]), the Republic of Congo (RoC, n = 22 [1.35%]), and Cameroon (CAM, n = 2[0.12%]). To this broad geographic sampling, was added a set of contextual clade Ia genomes from DRC (26), CAR (10), Cameroon (2), Gabon (2), South - Sudan (2) and Republic of Congo (1).

Phylogenetic analysis classified the genomes into different main clusters, defined arbitrarily as groups (Figure 2A). Among the previously described Clade Ia groups, Group I included genomes from Gabon (2), Cameroon (4), and the Central African Republic (CAR) (2). The two CAR genomes specifically originated from districts near Cameroon’s border. Notably, this group excluded genomes from the Democratic Republic of the Congo (DRC) or the Republic of the Congo (RoC), underscoring the virus’s circulation within a distinct regional reservoir.

**Figure 2.**
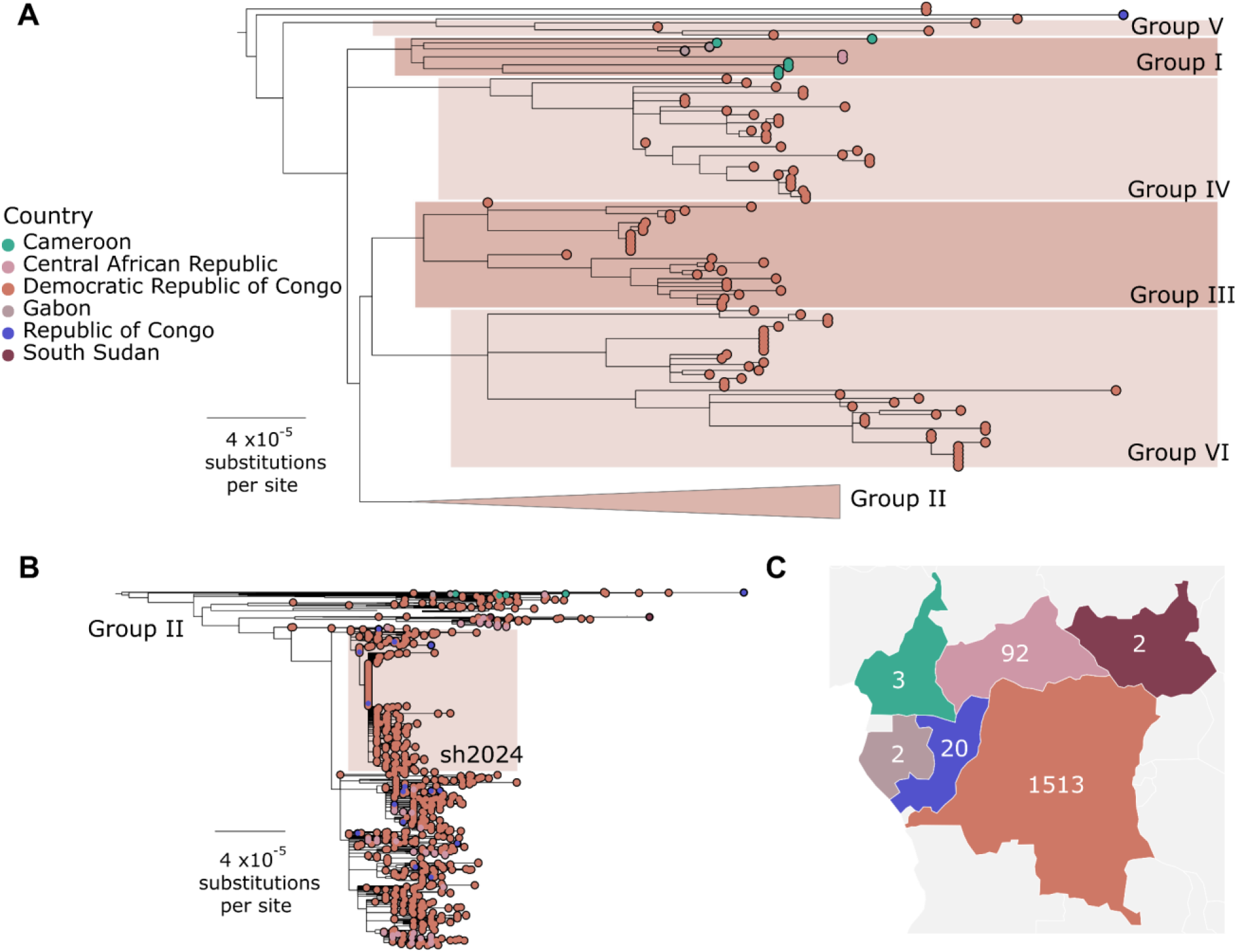
Maximum likelihood phylogenetic tree of MPXV virus Clade Ia. Panel A: Overall tree with Group II genomes collapsed, providing a broad view of Clade Ia diversity. Panel B: focused Group II subtree, highlighting its internal structure and sub-clustering.

Group II emerged as the dominant lineage, encompassing 1,536 genomes. This group included 20 RoC genomes, 90 from CAR, and 1,426 from DRC. Group II not only harboured the largest number of genomes but also exhibited the greatest genetic diversity (Figure 2B). Interestingly, Group II also included genomes related to the sh2024 outbreak that emerged from Kinshasa, the DRC capital. Group III comprised 27 genomes, all originating from the Democratic Republic of Congo (DRC). Within this group, two distinct subclusters were identified with 13 and 14 genomes, respectively. Additionally, we identified a novel lineage, noted ad group VI closely related to Group III, consisting of 40 DRC genomes. Groups IV and V included 25 and 6 genomes, respectively, all of which originated from the DRC. These groups, though smaller in size, contributed to the overall phylogenetic landscape and highlighted the genomic complexity within the DRC. Finally, 2 genomes from the DRC and 1 from RoC did not cluster with any of the above-described groups, suggesting the potential existence of other groups, as it was the case for Group V.

### Genomic diversity of the MPXV Clade Ib

A total of 1,305 genome sequences belonging to Clade Ib were obtained from 10 AU Member States, with Uganda and the Democratic Republic of Congo having the highest number of genomes. The samples were collected between December 2023 and June 2025, with the DRC exhibiting the longest sampling period.

At the root of the phylogenetic tree are genomes derived from the Democratic Republic of Congo (Figure 3). Three major clusters appear from the tree: Cluster 1 encompasses genomes from Uganda, Burundi, and Zambia; Cluster 2 includes genomes from Tanzania, Zambia, Uganda, DRC, Burundi, Ethiopia, and Kenya; and Cluster 3 consists of genomes that are more heterogeneous from Zambia, Kenya, the Republic of Congo, Burundi, Uganda, Malawi, DRC, and Angola.

**Figure 3.**
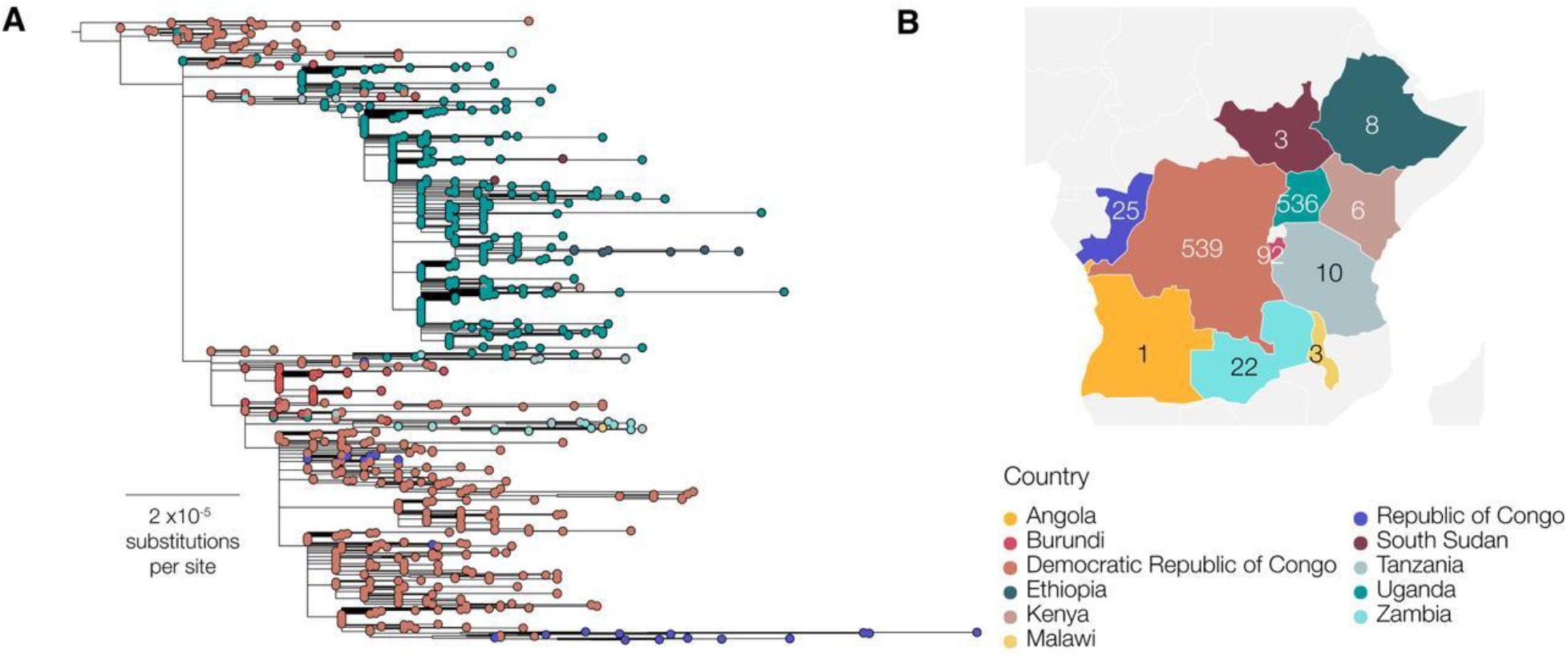
Phylogenetic structure of MPXV Clade Ib genomes across 10 African Union Member States. Genomes from the Democratic Republic of Congo form the basal lineage, with three major clusters emerging: Cluster 1 (Uganda, Burundi, Zambia), Cluster 2 (Tanzania, Zambia, Uganda, DRC, Burundi, Ethiopia, Kenya), and Cluster 3 (Zambia, Kenya, Republic of the Congo, Burundi, Uganda, DRC, Angola), reflecting regional diversification and lineage mixing.

Our results indicate that Subclade Ib of MPXV Clade I, initially identified in the Democratic Republic of the Congo (DRC) in 2011, has followed a distinct cyclical pattern of emergence, disappearance, and re-emergence across Africa, with genetically divergent strains documented in 2011–2012 and 2023–2025.

### Genomic diversity of the MPXV Clade IIa

The Clade IIa exhibited country-linked clusters consistent with predominantly localised transmission during 2017–2018 (Figure 4). Several short branches connected these clusters to European travel-associated tips—most notably a tightly clustered Sierra Leonean genome with a Netherlands genome, forming a short-branched, that suggests a recent epidemiologic link. Liberian and Côte d’Ivoire sequences formed clusters with short internal branches and limited within-country divergence; in some instances, their tips connected with neighbouring West African genomes, reflecting regional connectivity rather than independent sustained lineages.

**Figure 4.**
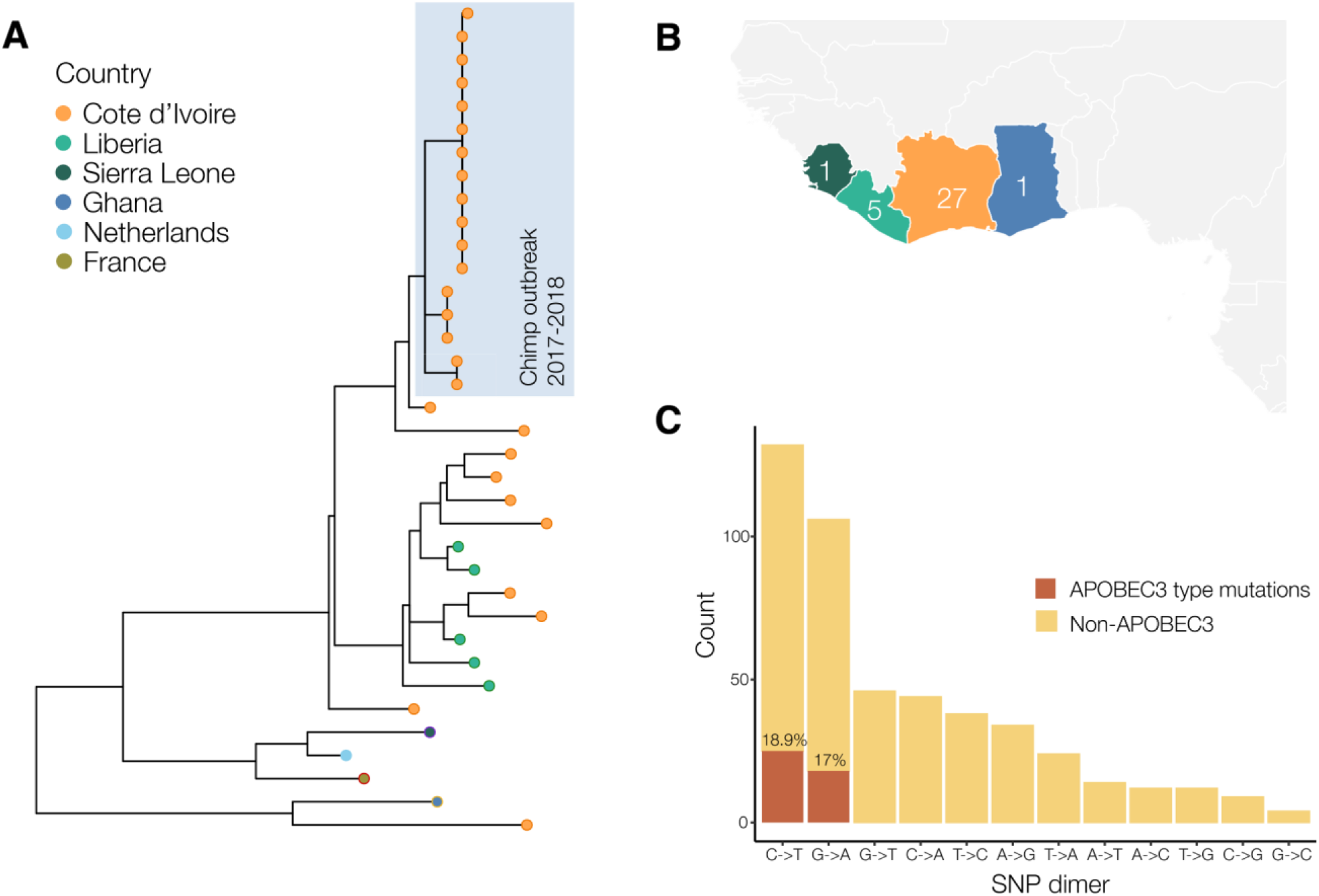
Phylogeny of monkeypox virus clade IIa genomes, West Africa. Ancestral state reconstruction of the clade II phylogeny, comparing the new sequences from the recent outbreak against all publicly available clade IIa genomes.

### Genomic diversity of the MPXV Clade IIb

This combined analysis initiative has also shed light on the dynamics of Clade IIb. The first findings revealed the concurrence of both zoonotic and human-to-human (H2H) transmission driven by MPXV clade IIb strains across affected countries in Africa. We found that all 17 sequences sampled between 2018 and 2022 in Cameroon form a divergent basal sister lineage to hMPXV-1, and its zoonotic outgroup from Nigeria. The zoonotic cluster also included sequences that originated from a bordering state in Nigeria. There is limited evidence of APOBEC3-mutation enrichment (% apobec3 rate) in this cluster, confirming the likely zoonotic origin of MPXV diversity circulating in the Cameroon-Nigeria cross-border area. In the context of the 2024-2025 clade IIb outbreak in other affected countries, our analysis revealed that the sampled cases were classified as lineage A.2.2 and exhibited signatures of H2H transmission, exhibiting enrichment of APOBEC3-type mutations. Specifically, the frequency of G→A and C→T mutations reached 90.2% and 92%, respectively, a dramatic rise compared to the 16% and 18% Nigeria and Cameroon zoonotic cases from samples collected between 2017-2023. In Sierra Leone, the genomic data revealed low intra-country diversity, indicative of active local transmission rather than multiple introductions (Figure 5). This was further corroborated by global phylogenetic mapping, which traced the exportation of lineage A.2.2 (G.1) from Sierra Leone to Germany & USA (Figure 5). In Togo, the analysis allowed to document another strain descending from the A.2.2 lineage (here called G.2).

**Figure 5.**
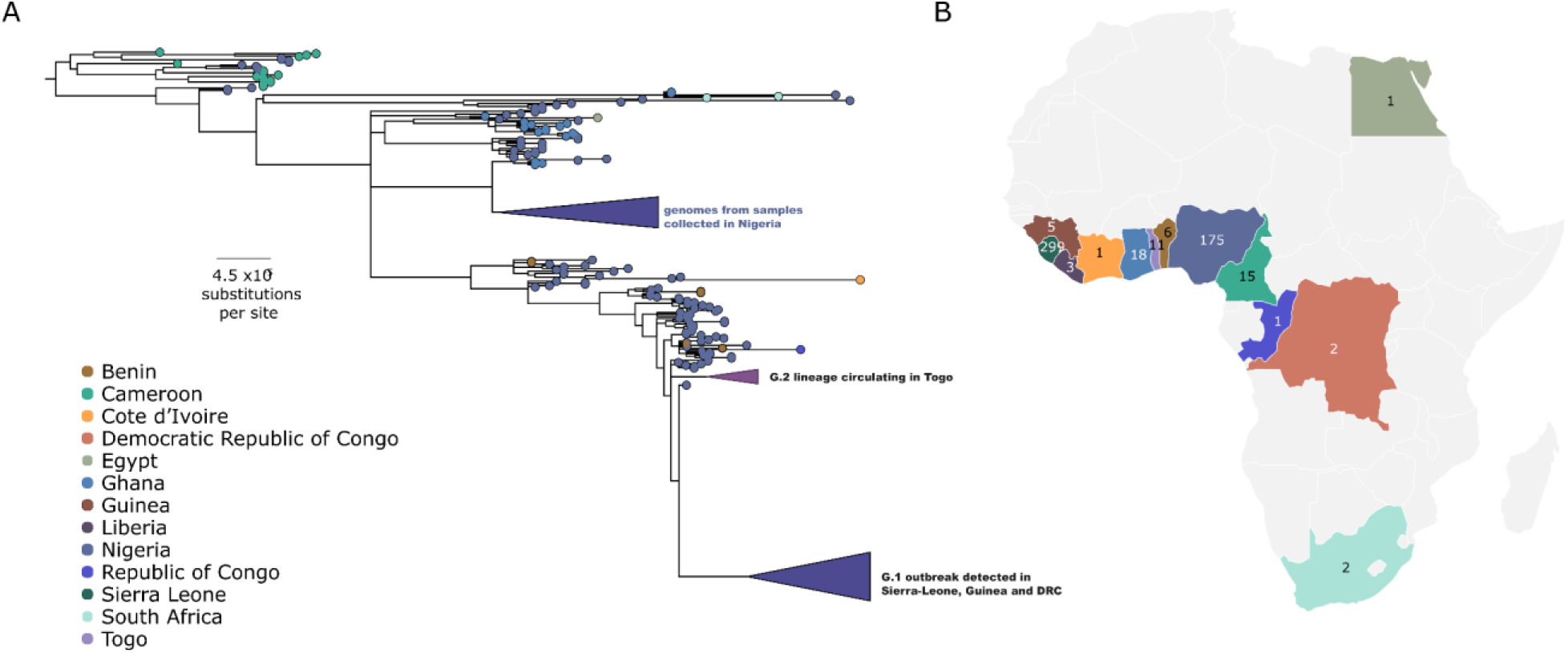
Transmission modes and evolutionary signatures of MPXV Clade IIb across Africa. Genomic analyses reveal a divergent zoonotic lineage at the Cameroon–Nigeria border characterized by minimal APOBEC3 activity. In contrast, lineage A.2.2 shows widespread H2H transmission and elevated APOBEC3-type mutations, highlighting its regional spread and adaptive evolution.

In contrast, Nigeria and Benin presented a more complex picture. Multiple distinct clusters were identified, pointing to several independent introductions of Clade IIb. Notably, one Nigerian genome aligned with the 2022 zoonotic outbreak, suggesting ongoing spillover events alongside human transmission. So far, lineages A, B and G have been identified in African countries.

## Discussion

The recent MPXV epidemic in Africa underscores the increasingly complex and dynamic nature of MPXV transmission ^22–24^. Our genomic analysis of more than 3,000 high-quality genomes from 24 AU Member States demonstrates that multiple clades (Ia, Ib, IIa, IIb) are co-circulating across the continent.

These diverse clades exhibit distinct evolutionary trajectories and epidemiological patterns, contributing to worsening health outcomes, the rapid spread, and difficulties in containment and prevention. With different genetic makeup, clades like the more virulent Clade I cause severe illness and have higher mortality rates, while other clades can have milder symptoms but are still highly transmissible ^6,7,25,26^. The heterogeneity observed reflects both ecological diversity in animal reservoirs and the intensifying role of H2H transmission, particularly within key populations and across porous borders. However, the shift to sustained human transmission would likely be attributable to epidemiological factors, rather than an intrinsic increase in the virus’s transmissibility or virulence ^27,28^.

This diversity necessitates complex, multi-faceted public health responses, including surveillance, targeted vaccination, and robust public health guidance to mitigate the varying transmission patterns and clinical manifestations across different clades and regions ^29,30^.

### Clade Diversity and Evolutionary Patterns

These findings confirm previously reported clade structures while revealing novel insights into intra-clade diversity. Clade Ia appears predominantly reservoir-driven across Central Africa, with the DRC, RoC and CAR acting as core areas of zoonotic maintenance and diversification. The DRC in particular harbours deep intra-clade diversity (Groups II-V) and potential novel groups (VI), a DRC-specific cluster, consistent with long-term viral maintenance and local evolution. Neighbouring interfaces such as Cameroon and Gabon, Cameroon and CAR, DRC and CAR or DRC and RoC provide clear examples of continued zoonotic spill-over and cross-border movements. Moreover, the first documentation of CAR genomes within Group I-predominantly composed of Gabon and Cameroon genomes-demonstrates how expanded sampling enhances our understanding of the virus’s spatial distribution and reinforces the importance of broader regional genomic surveillance. However, we also observe the emergence of sustained H2H strains within the DRC, indicating that zoonotic maintenance and human transmission now coexist within this clade Ia ^7,13^. Genomic patterns, together with transmission in key populations and mobility linked to insecurity and rural vulnerability, drive local amplification and regional spread, intensifying the public health threat beyond traditional hotspots.

Unlike clade Ia and the ancestral clade I, clade Ib demonstrated sustained H2H transmission, suggesting the involvement of cryptic reservoirs or undetected asymptomatic infections. Its emergence was first observed in the DRC through clustered outbreaks in the eastern region, before spreading to neighbouring countries, where large epidemics were recorded in Uganda and Burundi ^7,13,31^. Multiple introductions have since been detected in other non-endemic countries across Eastern and Southern Africa, including Ethiopia, Kenya, Tanzania, and Malawi ^32,33^, underscoring its expanding geographic reach and epidemic potential.

Clade IIa cases still occur in West African countries, including Sierra Leone, Ghana, Liberia, and Côte d’Ivoire, where it has circulated for decades through sporadic zoonotic spillovers. Although historically associated with milder disease and lower-case fatality rates (<1%) compared to Clade I ^34^, Clade IIa has a persistent transmission potential, with occasional expansion ^5^. Though less well represented, it appears to sustain both zoonotic introductions and limited H2H spread, consistent with observations from Côte d’Ivoire in 2024 ^35^. Despite its relatively stable epidemiological profile, the continued circulation of Clade IIa in ecologically vulnerable zones and its genetic divergence from other clades do not preclude the evolution of more adapted strains that could emerge, underscoring the need for targeted surveillance and strengthened public health preparedness

Clade IIb shows prolonged human transmission, with lineage A.2.2 showing signatures of APOBEC3 mutational activity (>90%), confirming sustained H2H spread and accelerated viral evolution ^36,37^. At the Cameroon-Nigeria border, divergent lineages with minimal APOBEC3 signatures confirmed repeated zoonotic spillovers, while Nigeria emerged as both a site of recurrent introductions and a hub of ongoing transmission ^22^. In Sierra Leone, limited intra-country diversity suggested localised transmission, whereas the detection of lineage A.2.2 in Germany underscored the potential for international spread ^9,38^. This pattern is best interpreted as a two-step evolutionary dynamic: repeated zoonotic introductions continually seed genetic diversity, and when those variants enter dense human networks, APOBEC3-associated editing and selection accelerate within-human evolution of the virus population, thereby increasing the lineage’s epidemic potential. Together, these findings show that Clade IIb outbreaks are driven by both ecological spillovers and human transmission, emphasizing the need for cross-border genomic surveillance and its rapid integration into public health action.

### Cross-Border Transmissions

Our near continent-wide dataset reveals that MPXV transmission in Africa is fundamentally regional: viral lineages routinely cross-national boundaries and follow human movement, trade routes and displacement corridors rather than political borders. The DRC emerges as a major reservoir and diversification hub, seeding multiple intra-clade lineages (Clade Ia Groups II–V and novel DRC-specific clusters) that have dispersed to neighbouring countries (Uganda, Kenya, and Burundi); these DRC-centred diversifications underpin much of the cross-border signal observed in the region. By contrast, the Cameroon–Nigeria axis illustrates a predominantly zoonotic cross-border interface: fifteen Cameroonian sequences sampled between 2018–2022 form a divergent, basal sister lineage to hMPXV-1 and cluster with genomes from a bordering Nigerian state, and this zoonotic cluster displays a very low APOBEC3 editing signature consistent with non-human origin. West Africa shows a distinct, heterogeneous pattern in which Nigeria and Benin harbour multiple independent Clade IIb introductions indicative of repeated spillovers and local seeding, whereas Sierra Leone demonstrates low intra-country diversity consistent with local expansion from a single introduction and documented exportation of G.1 lineage (for example, travel-associated exportation to Germany) ^9,38^.

These phylogeographic patterns are mirrored by molecular signatures: where variants enter dense human networks—most strikingly lineage A.2.2 in the 2024–2025 outbreak—APOBEC3-associated G→A/C→T enrichment rises sharply (G→A and C→T frequencies of ∼90% and ∼92% in 2024–2025 versus ∼8.6% and 22.6% in 2022–2023), producing a clear genomic footprint of prolonged human chains and elevated epidemic potential. Taken together, the geographic and molecular evidence support a two-step model of regional spread in which ecologically driven spillovers generate the standing diversity, human mobility and porous borders disseminate that diversity across subregions, and concentrated transmission networks in receiving locales amplify onward spread. These findings therefore argue strongly for transnational, harmonised genomic surveillance and coordinated cross-border response mechanisms—including joint One Health investigations at known interface zones (e.g., Cameroon–Nigeria and DRC border areas), routine APOBEC3-aware genomic triage, and rapid shared epidemiological follow-up—to ensure that local spillovers do not become propagated regional epidemics.

### Implications for Public Health and Genomic Surveillance

The co-circulation of multiple MPXV clades capable of sustained H2H transmission poses an urgent threat to public health security in Africa and globally. Recombination signals within Clade Ib, together with APOBEC3-driven evolution in Clade IIb, reveal the virus’s extraordinary evolutionary potential to generate fitter, more transmissible lineages ^6,7,39^. Without early detection and rapid response, these changes risk accelerating epidemic spread, particularly in health systems that are already stretched.

The continental analysis of MPVX sequencing data marked a major milestone for public health in Africa, demonstrating unprecedented collaboration among AU member states. This coordinated effort not only mapped the virus’s evolution and transmission across the continent with unprecedented precision but also overcame traditional barriers to data sharing. By pooling resources and expertise for shared analysis, this work set a powerful precedent for the collective management of future epidemics. It now serves as a model for establishing more robust, equitable, and effective data-sharing frameworks, ensuring that crucial genomic information can be rapidly mobilized for public health decision-making at regional and global levels.

However, major surveillance blind spots remain. Persistent gaps in genomic sequencing and reporting— most notably in Central and Eastern Africa—undermine our collective ability to identify introductions, trace transmission, and mount timely interventions. This inequity in surveillance capacity and data sharing leaves entire regions vulnerable to unchecked spread, delaying the global response and magnifying epidemic risk.

Strengthening Africa’s genomic surveillance architecture is urgent, but it cannot stand alone. Vaccines must be made rapidly accessible, affordable, and equitably distributed across affected countries. Without vaccination as a frontline tool, surveillance alone will not curb ongoing transmission or prevent further regional and international dissemination. Integrating real-time genomic surveillance with widespread vaccination, timely clinical management, epidemiological intelligence, and cross-border coordination represents the strongest pathway to containment.

MPXV now represents both a regional and global health security challenge. Closing surveillance gaps, investing in equitable sequencing capacity, and ensuring timely vaccine access are not optional; they are essential pillars for curbing this epidemic and for strengthening preparedness against the next emerging pathogen. To contain MPXV effectively, a holistic One Health approach is urgently needed, alongside intensified human case detection, programmes must include systematic sampling of rodents and other suspected reservoir species to clarify zoonotic spillover dynamics; targeted environmental surveillance of high-risk settings (markets, farms and bushmeat hubs) at human–animal interfaces; and population-level seroprevalence studies to capture symptomatic and asymptomatic infections and define prevalence, incidence and immunity gaps.

To translate these findings into an actionable public health impact, we propose a targeted regional strategy designed to enhance surveillance, guide interventions, and strengthen cross-border coordination. First, expand representative sequencing into geographically and ecologically diverse sentinel sites and mandate harmonised minimum metadata (travel, exposure, network membership, clinical severity) with every sequence; concurrently, operationalise APOBEC3-based genomic triage so that sequences showing human-specific signatures automatically prompt epidemiological investigation and escalation of control measures. Second, recognise MPXV as a regional problem and build transnational surveillance networks and cross-border rapid-response teams that link joint One Health investigations at interface zones (coordinated animal sampling paired with human genomics) to real-time genomic outputs. Third, invest in durable regional capacity: harmonised wet-lab and bioinformatics SOPs, external QA, sustained training, and mobile surge teams, and routinely integrate genomic data with serosurveys, mobility/displacement metrics and environmental surveillance (e.g., wastewater/xenosurveillance). All technical measures should be underpinned by clear ethical, legal and data-sharing frameworks to protect privacy, promote timely information exchange, and ensure community engagement. Implemented together, these actions will enable genomics to drive rapid, proportionate, and coordinated public-health responses to MPXV across the continent.

Following recommendations from the Africa CDC Emergency Consultative Group, mpox was formally lifted as a PHECS, marking a transition from emergency response to sustained, country-led control and elimination strategies. Importantly, this transition does not signify the end of mpox transmission in Africa. Rather, it underscores the need to institutionalise genomic surveillance, maintain cross-border coordination, and apply integrated genomic-epidemiological approaches to detect resurgence, guide interventions, and inform diagnostics and vaccine development.

## Methods

### Study Design, Data Collection, and Preprocessing

For our study, we conducted a continent-wide genomic analysis of MPXV in Africa, integrating 3,450 genomes alongside publicly available sequences representing Clades Ia, Ib, IIa, and IIb, spanning the period from 1970 to 2025, from over 24 African Union Member States. All Member States voluntarily contributed their Mpox genomic data through a secure, cloud-based Terra platform, enabling integrated analyses, harmonized interpretation, and the development of a unified, continent-wide study. This is a cross-sectional and retrospective study based on a collection of newly complete and near-complete MPXV sequences. Publicly available sequences were retrieved from open-access databases such as the Global Initiative on Sharing All Influenza Data (GISAID) and other curated repositories (Pathoplexus), as of 21 August 2025. Different sequencing approaches and platforms were used after the confirmation of the cases by RT PCR. The study included only the sequences associated with minimum demographic metadata, such as the date of sample collection, the country and the region.

Genome Assembly: Samples from 24 AU Member States, genome assembly was conducted depending on the library preparation strategy. For data generated through probes hybridization capture approach, the genomes were assembled using different pipelines such as the viral-ngs assemble_denovo_metagenomic pipeline with automated reference genome selection, czid, consensus fasta, metatropics or other in-house country-specific pipelines. When amplicon sequencing was used as an enrichment approach, genomes were assembled using the ARTIC-MPXV pipelines, either for Illumina or Nanopore, respectively.

Phylogenetics: We used NextClade v3.16.0 ^40^ to assign clade and lineage to each genome according to the nomenclature ^41^. This clade assignment was used to segregate the genomes into clade-specific analysis. Genomes were compiled for each clade, with accompanying metadata of country of origin and collection date. For each clade, we used Squirrel v1.2.2 to perform alignment, phylogenetics and APOBEC3-reconstruction ^42^. Squirrel aligns MPXV genomes by mapping genomes against either the Clade I or Clade II RefSeq (Accession numbers: NC_003310 and NC_063383, respectively) using minimap2 and constructs an alignment using gofasta ^43,44^. By default, the 3’ inverted terminal repeat region and a set of known problematic regions are masked. It automatically selects an appropriate outgroup to root the tree based on the clade specified and uses IQTREE2 to estimate a maximum likelihood phylogeny, using the HKY model^45^. Ancestral state reconstruction is run, and Squirrel uses these node state reconstructions to infer the mutations that occurred along each branch in the phylogeny. Squirrel categorises each SNP by whether it appears in an APOBEC3 context or not and plots a phylogeny with those reconstructions along the branches.

Initially, SQUIRREL was run in Quality Control (QC) mode, which flags SNPs that may be derived from assembly or alignment issues (e.g. clustered SNPs, N-adjacent SNPs, convergent SNPs or reversions to reference. Each of these was investigated and inspected by eye, and then Squirrel was run again with an additional mask file that included the flagged clustered and N-adjacent SNPs. The convergent SNPs were not masked, and the reversions were used to flag potential assembly errors in the genomes.

For the combined phylogenetic analysis, a subset of genomes was selected that represented the major diversity of each clade (Total genomes 218, and 64, 100, 13 and 41 for Clades Ia, Ib, IIa, and IIb, respectively). Clade I and Clade II genomes were first aligned and masked separately using SQUIRREL, against their respective RefSeq’s and masked using the appropriate mask files, then the alignments were combined using mafft-profile alignment, which assumes phylogenetic independence for each clade ^46^. IQTREE2 was used to estimate a maximum likelihood tree, with the HKY model and 1000 ultrafast bootstraps ^45^.

## Data Availability

All data produced in the present study are available upon reasonable request to the authors

## Ethical Statement

This study analysed de-identified surveillance data collected for public health purposes. The Mpox affected African Union (AU) Member States collaborated by sharing data in a secure joint space on Terra.bio (https://app.terra.bio/#workspaces/gates-pgs-africacdc/Africa%20PGI%20Mpox) and met twice to conduct a joint bioinformatic analysis. The participating AU member states collaboratively developed the current manuscript, interpreting the results under the guidance and supervision of senior leadership. All authors representing the participating AU member states approved the final version for publication. Because the dataset was fully anonymised and no direct interaction with human participants occurred, the requirement for informed consent was waived. Where applicable and in accordance with local regulatory policies, ethical approval was either obtained or formally waived to enable the use of the data.

The following countries had ethical clearance included: **Burundi:** National Ethics Committee in Burundi (CNE/10/2024); **Cameroon:** Cameroon National Ethics Committee for Research in Human Health (Approval Number: 2020/05/1224/CE/CNERSH/SP); **Central Africa Republic (CAR):** Ethics and Scientific Committee/University of Bangui (Approval Number: N°33/UB/FACSS/IPB/CES/024); **Democratic Republic of Congo (DRC):** Ethics committee of the Kinshasa School of Public Health (Kinshasa, Congo; approval number ESP/CE/238/2024); **The Republic of the Congo (RoC):** The Congolese Foundation of Medical Research Ethics Committee (Approval Number: 053/CEI/FCRM/2024); **South Sudan:** Ministry of Health Research and Ethics Review Board (MOH-RERB); **United Republic of Tanzania:** National Health research ethics committee (NathREC); Uganda: National Health Laboratory Services Research and Ethics Committee (UNHLS-2025-133).

The following countries had ethical clearance waived included or data already available on public repositories (Genbank, GISAID and/or Pathoplexus): Angola, Benin, Cote d’Ivoire, Egypt, Ethiopia, Gabon, Ghana, Guinea, Kenya, Liberia, Malawi, Morooco, Nigeria, Sierra Leone, South Africa, Togo, Zambia.

## Acknowledgements

We gratefully acknowledge all data contributors, including the authors, their originating laboratories and partners for sharing MPXV sequences and metadata. We also acknowledge the support of all national broad teams and international consortia and agencies that have contributed to the generation of data. Finally, thanks to Egypt, France and the Netherlands for the contextual genomes they previously made available in public repositories

## CRediT author statement

Conceptualized the manuscript: CKT, SKT, IS, PMK, and YKJ. Methodology: CKT, EKL, MC, AOT, AR, AKC, MMD and SK. Software: AOT and AR, Validation: JW, HH, RC, HAK, EN, AS, FRN, RN, SN, IS, EL, GN, GI, OP, FS, AA, PMK, SKT, YKJ. Formal analysis, Investigation, and Data curation: All Authors. Writing – original draft: CKT, EKL, ACK, MMD, JHK, and RN. Writing – review & editing: All Authors. Visualization: AOT, MC, EKL, MMD, AR, MG, and ELL. Supervision: AR, IS, PMK, SKT, and YKJ. All authors have approved the final version of this manuscript for submission for publication.

